# A Tip Optofluidic Immunoassay System for COVID-19 Immuno-protection Evaluation Using Fingertip Blood

**DOI:** 10.1101/2024.04.29.24306554

**Authors:** Ruihan Li, Binmao Zhang, Hao Li, Lixiang An, Tianen Zhu, Shi Hu, Fan Yang, Weishu Wu, Xudong Fan, Yujuan Chai, Hui Yang, Xiaotian Tan

**Affiliations:** Institute of Biomedical and Health Engineering, Shenzhen Institute of Advanced Science, Chinese Academy of Science, Shenzhen 518055, China; The Key Laboratory of Biomedical Imaging Science and System, Chinese Academy of Sciences, Shenzhen 518055, China; Department of Biomedical Engineering, Shenzhen University Medical School, Shenzhen University, Shenzhen 518060, China; National Innovation Center for Advanced Medical Devices, Shenzhen 518110, China; Emergency Department, Shenzhen University General Hospital, Shenzhen University, Shenzhen 518060, China; Department of Biomedical Engineering, University of Michigan, MI 48109, USA

**Keywords:** Immunoassay, Optofluidics, Antibody Assessment, COVID-19, Fingertip Blood

## Abstract

Infectious diseases such as COVID-19 continue posing significant global health challenges, with recurrent re-infections contributing to long-term symptoms such as cardiac issues and anosmia. Effective management of re-infections relies heavily on maintaining high levels of circulating binding and neutralizing antibodies. Traditional methods for antibody quantification, such as ELISA, face significant challenges, including narrow dynamic ranges and complex sample preparation procedures, which hinder their applications in rapid and routine diagnosis. This study introduces a novel optofluidic biosensing technology, tip optofluidic immunoassay (TOI), that addresses these limitations by enabling the quantitative analysis of binding IgG against multiple SARS-CoV-2 strains from only 1 μL of fingertip blood. The proposed TOI system, featuring industrial-grade micro-fabricated immuno-reactors and a portable chemiluminescent imaging station, can provide test results within 12 minutes. For IgG binding assays, TOI possesses a lower limit of detection of 0.1 ng/mL, a dynamic range of 3-4 orders of magnitude, along with a high signal-to-noise ratio (approximately 10,000). This technology not only simplifies the antibody quantification process but also enhances patient compliance and facilitates decentralized testing, which is crucial for infectious disease management. By enabling precise and rapid antibody assessment, this system can support the optimization of vaccination strategies and broader public health responses to COVID-19 and other infectious diseases.

## Introduction

The COVID-19 pandemic has cast a long shadow worldwide for the past four years, posing a continuous threat to public health^1, 2^. Research suggests that recurrent re-infections of SARS-CoV-2 variants could elevate the risk of long-COVID symptoms, including fragility, sleep disturbances, cardiac issues, and anosmia, severely affecting one’s quality of life^3, 4^. One of the crucial elements in reducing COVID-19 re-infection is to maintain a substantial concentration of circulating neutralizing antibodies^5, 6^. However, given the diverse COVID-19 vaccinations administered, the variance in strains infected, and the inoculation times across the population, pinpointing a uniform immune background remains an impossible goal^7, 8^. Consequently, the accurate assessment of binding and neutralizing antibody levels emerges as a pivotal factor in deciding the optimal timing and choice of vaccine boosters^9, 10^.

Unfortunately, current techniques for the quantitative evaluation of SARS-CoV-2 binding and neutralizing antibodies failed to become a standard examination in routine disease control operations, primarily due to technical limitations in rapid assay methodologies^11, 12^. Traditional methods like ELISA face obstacles for accurate antibody quantification, attributable to their narrow dynamic range (∼1.5 orders of magnitude)^13^. The concept of “titer” commonly refers to antibody concentration, yet its determination involves complex dilution processes. Such assays not only are labor intensive, sample wasting (venous blood ∼ 1 mL), but also render cross-platform comparisons challenging due to susceptibility to methodological and environmental variations^14^.

Fingertip blood immunoassays represent a new concept approach in diagnostics and are getting popular among the population by offering the convenience and minimally invasive sample collection for rapid testing^15–17^. This method significantly enhances patient compliance and facilitates decentralized testing, which is crucial for infectious disease management^18, 19^. Micro-volume samples such as the fingertip blood offers a “perfect match” with microfluidic biosensors^20–23^. However, the integration with microfluidic technologies still confronts substantial hurdles. Most of the existing microfluidic biosensing assays, despite their potential for precision and portability, often grapple with the complex composition of whole blood^24, 25^, leading to the issues like microchannel clogging and interference with the biochemical reactions^26^. Furthermore, the challenge of maintaining the stability and functionality of reagents within the complicated and confined channels of microfluidic devices weakens their ability to provide accurate, sensitive, and specific results^27, 28^. Except for a few exceptions, most of the microfluidic immunoassays did not consider the significance of neutralizing antibody evaluation, thus reducing the utility of their assays^29, 30^.

Here, we are introducing a new member of the optofluidic biosensor family: the tip optofluidic immunoassay (TOI) system. This in-house developed technology allows for quantitative and comprehensive immune assessment of SARS-CoV-2 (including multiple strains) binding IgG using just 1 μL of fingertip blood. The proprietary TOI system features industrial-grade micro-fabricated and mass-produced microfluidic immuno-reactors that can be integrated with pipette tips, along with a custom-designed portable chemiluminescent imaging station and a liquid handling system, forming the core hardware of this technology. Our optimized immunoassays not only quantify SARS-CoV-2 binding antibody levels within 12 minutes, with a lower limit of detection (LLOD) of 0.1 ng/mL and a dynamic range spanning 3-4 orders of magnitude, but also boast an exceptionally high signal-to-noise ratio (∼10000). This work offers a streamlined avenue for assessing antibody-mediated immuno-protection against COVID-19, with significant implications for vaccine development and managing other infectious diseases in the foreseeable future.

## Materials and Methods

### Portable Tip Optofluidic Immunoassay System (TOI)

The TOI system is composed mainly of two sections: (1) microfluidic immuno-reactors and (2) portable chemiluminescent imaging station. The immuno-reactors are used as disposable structures for conducting immunosorbent assays with fingertip blood samples; the chemiluminescent imaging station is designed to quantitatively measure the chemiluminescent signal intensities of the immuno-reactors. Detailed information for the two sections can be found below:

### Microfluidic immuno-reactors

The microfluidic immuno-reactors were fabricated through injection molding with high-protein affinity polystyrene, with technical support from Beijing 4.0 Industrial Technology Co., Ltd. The microfluidic immuno-reactors are composed of three units: sensing unit, intersection, and adapting unit. The inner diameter of the microfluidic immuno-reactors is 0.9 mm. As presented in Fig. S1, the industrial-grade mass microfabrication of the reactors could achieve a qualified yield of over 99.9 % for each batch of product (∼10000), indicating a high reliability. To perform an immunoassay, the microfluidic immuno-reactors will be connected to standard 20-200 μL pipette tips for liquid handling. No chemical surface treatment is needed before each immunoassay. The evaluation of protein immobilization kinetics for the microfluidic immuno-reactors is shown in Figure S2A. For common proteins such as IgG, physical adsorption generally saturates after 20-40 minutes, which is significantly faster than conventional immunoassay reactors like 96-well plates.

### Chemiluminescent imaging station

To simplify the platform structure to the greatest extent, the chemiluminescence imaging method was chosen for immunoassay signal measurement. The portable chemiluminescent imaging station was designed and assembled in-house. As presented in Fig. 1, The chemiluminescent imaging station includes a shading cover (stainless steel), a pipette holder (also works as the immuno-reactors), and a CMOS camera. The total weight of the signal measurement box is about 3 kg, which is easy to carry with one hand.

**Figure. 1.**
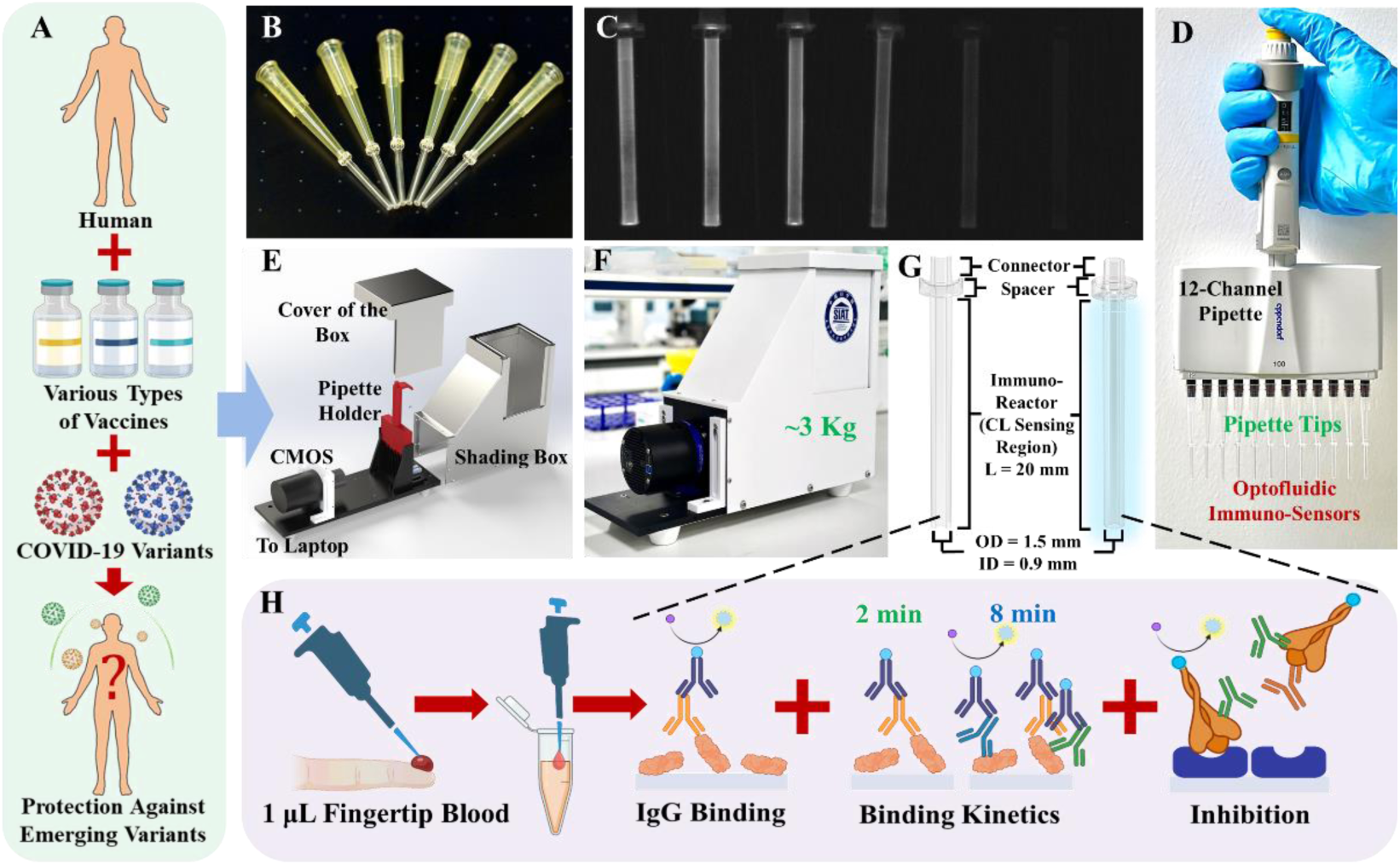
Concept of COVID-19 immuno-protection evaluation using TOI. (A)Different scenarios stemming from varied vaccination and infection histories can complicate the evaluation of immuno-protection efficacy against emerging COVID-19 variants for the general public. (B) Key components of the TOI system: polystyrene microfluidic immuno-reactors connected with pipette tips. (C) Chemiluminescence images of the immuno-reactors. (D) Photo of the immuno-reactors under ambient light operated with a 12-channel pipette. (E) Structural illustration of the portable chemiluminescent imaging station (PCIS), with a CMOS camera as the chemiluminescent signal detector. (F) Photo of the TOI PCIS prototype. (G) Schematic of batch-producible polystyrene immuno-reactors. (H) Workflow for SARS-CoV-2 immuno-protection evaluation, requiring only a microliter of fingertip blood to assess IgG binding, kinetics, and neutralization capacity.

The intensity of the chemiluminescence signal is generally very weak. Thus, the protection against environmental light interference is a key factor in the design of the signal measurement box. Light-protection structures are placed in all corners and connecting sections. To perform accurate and quantitative measurements of the chemiluminescent system, we used a highly-sensitive monochromic CMOS camera QHY533M (from QHYCCD) along with a Fujinon wide-angle micro-distance lens to collect long exposure chemiluminescent images. The camera is placed 12.5 cm in front of the pipette holder. The pipette holder (fabricated through 3D printing) is designed to stabilize the multi-channel pipette and the attached immuno-reactors attached. Vertical walls are built between the microfluidic immuno-reactors to prevent inter-channel crosstalk.

The optical quantification performance of the QHY533M camera can be found in Fig. S2(B). The minimum detectable optical signal intensity was 1.0, and the linear dynamic range was about 60,000 (at gain = 70), covering 4.5-5 orders of magnitude in optical signal intensity.

### Biomolecular and Chemical Reagents

All fundamental chemical reagents, including 10 × PBS buffer, pH=7.4 (AM9624), wash buffer (0.05% Tween-20 in PBS), and luminol chemiluminescence substrate (SuperSignal^TM^ ELISA Femto Substrate, 37075) were purchased from Thermo Fisher. All protein buffers, including 10% Blocker^TM^ BSA (in PBS, 37525), 1% casein (in PBS, poly-HRP dilution buffer, N500), and SuperBlock^TM^ buffer (in PBS, 37515) were also purchased from Thermo Fisher.

For biomolecular reagents, broad-spectrum SARS-CoV-2 antibodies SA55 and SA58 were developed and provided by Dr. Yunlong Cao from Peking University. Recombinant SARS-CoV-2 RBDs (40592-V08H for WT, 40592-V08H136 for XBB.1.16), recombinant SARS-CoV-2 S-ECD homotrimers (40589-V08H8 for WT, 40589-V08H48 for XBB.1.16), recombinant ACE2 (hFc Tag, 10108-H02H) and recombinant SARS-CoV-2 antibody D006 (40589-D006) were all produced and provided by Sino Biological. The AVI-tagged S-ECD homotrimers for WT and XBB.1.16 strains were designed, expressed, and purified by Sino Biological. The monoclonal anti-human-IgG antibody (HRP conjugated) was also developed and provided by Sino Biological.

### Human Sample Collection

All human samples, including fingertip blood, fingertip serum, venous blood, and venous serum were collected under SIAT-IRB-230715-H0667. Venous blood samples were collected by trained experts using vacuum blood collection tubes. Fingertip blood samples were collected with pain-free fingertip blood pens that were designed for routine blood glucose surveillance. Fingertip serum samples were generated by centrifuging 30 μL of fingertip blood for three minutes at 3000g. For all volunteers, the fingertip blood was diluted 25 times using 1% casein in PBS for storage. All other types of samples were also diluted with 1% casein in PBS before applying to TOI. The negative control sample was obtained from one of our previous clinical studies, which was approved by the IRB of the Second Affiliated Hospital of Chongqing Medical University (No. 2018 (100))^32^. The blood sample was collected before the COVID-19 outbreak and stored at −20 ℃ without freeze and thaw ever since. The informed consent of this study was obtained through re-contact of the healthy blood donor of the sample.

### Assay Protocol

For SARS-CoV-2 IgG binding assays, antigenic proteins (RBDs or S-ECD homotrimers) were first immobilized on the inner surface of the immuno-reactors. The working solution of protein was in 1 × PBS (pH = 7.4) at 10 μg/mL. Two consecutive blocking steps (3% BSA in PBS + SuperBlock^TM^ buffer) were used to reduce the noise level. The immuno-reactors were rinsed once after each incubation step. A graphical illustration of the assay protocol can be found in Fig. S3.

For concept demonstration, standard antibody solutions at different concentrations were tested with TOI. Three recombinant SARS-CoV-2 IgGs (SA55^33, 34^, SA58^35, 36^, D006^37^) were evaluated in this assay. The working solutions of the antibodies were first prepared by diluting the stock solution with 1% casein in PBS. Fourfold serial-diluted antibody solutions were drawn into the reactors to react with protein immobilized on the reactor’s inner surface and left at room temperature to react. The incubation time for the antibody solutions was 8 minutes. Subsequently, HRP-conjugated detection antibody solution (6000 × diluted with 1% casein in PBS, ∼170 ng/mL) was applied to the immuno-reactors. The incubation time for the detection antibody solutions was three minutes. Finally, a chemiluminescent substrate was introduced to the immuno-reactors after rinsing in triplicate. The optimization of the blocking and dilution buffer can be found in Fig. S4. Using 1% casein in PBS for both immuno-reactor blocking and sample/reagent dilution significantly reduced the background noise level to a nearly non-detectable level.

The IgG binding assay protocol for samples collected from participants is generally consistent with the proof-of-concept experiments. The only difference lies in the sample itself. As mentioned earlier, the original blood/serum samples were initially diluted 25× with 1% casein in PBS for storage. Various final concentrations can be prepared by further diluting the stock solution with 1% casein before applying to TOI. All other assay procedures are identical to the demonstration of concept experiments.

### Chemiluminescent Signal Quantification

After the final reaction step, the multichannel pipette with the microfluidic immuno-rectors is placed in a portable chemiluminescent imaging station to measure the chemiluminescence intensity. The baseline is calculated as the average blank signal intensity for the device. Based on our optimization results, 6 s exposure is generally suitable for most of the CL intensity quantification applications.

## Results

### TOI-based fingertip blood COVID-19 immuno-protection evaluation

In this study, a novel TOI system was developed to evaluate the immuno-protection efficiency of individuals with different vaccination histories, infection histories, and immune responses to SARS-CoV-2 variants (Fig. 1A). Considering that the immuno-protection assessment against various strains of infectious pathogens is a persisting task for public health, easy sampling, convenient operation, portable equipment, and low cost are of great importance.

The TOI system, as shown in Figures 1B-D, relies on the polystyrene microfluidic immuno-reactors that can be integrated with 200 μL pipette tips. The high surface-to-volume ratio of the reactors enables a fast and stable immobilization of concentrated biomaterials followed by efficient chemiluminescent immunoreactions in the capillary (Fig. 1C). With a 12-channel pipette, high throughput testing can be achieved even with manual operation. The signals will be detected using a 3 kg portable chemiluminescent imaging station (Figs. 1E-F), which can be carried out for field studies if necessary. Notably, the assay requires merely 1 μL fingertip blood (original sample volume) for quantitative comprehensive immune-protection assessments: the IgG binding test, the binding kinetics, and the rapid in-vitro inhibition assay (RIVIA) (Fig. 1H). These assays can be finished within 12 minutes while minimizing the risk and the cost. Generally, the multi-functional TOI system and all test procedures established fit the point-of-care needs of immuno-protection efficiency evaluation for emerging infectious diseases, providing a new solution to disease monitoring in clinical settings or decentralized scenarios.

### Establishment and evaluation of the IgG binding assays

With the TOI system developed, we first sought to establish the SARS-CoV-2 spike protein IgG binding assay using three renowned and well-calibrated monoclonal human IgGs, SA55, SA58, and D006, which have been verified in our previous studies ^38^. SA55 and SA58 were developed by Dr. Yunlong Cao and Dr. Xiaoliang Sunney Xie’s research group at Peking University. These two antibodies were proved to have broad-spectrum binding and neutralizing activities against almost all strains of SARS-CoV-2. Antibody 40150-D006 (abbreviated as D006) was originally developed against SARS-CoV-1 RBD by Sino Biological. The broad binding spectrum (but not neutralizing) against most SARS-CoV-2 strains was demonstrated in our previous studies^31^.

The SARS-CoV-2 RBD of two strains, WT and XBB was selected as the coating antigen on the microfluidic immuno-reactor. The former represents the ancestral strains that should be recognized by the immune system after the injection of inactivated vaccine, and the latter represents the new variants that are not likely under immuno-protection through the vast inoculation in China^39, 40^. We also designed and synthesized the trimeric versions of these two spike proteins for the IgG binding assays, mimicking the natural form of the protein and a more realistic interaction between the antigen and antibodies.

The calibration curves generated with chemiluminescent measurements can be found in Figures 2B-D. Note that for all results, the absolute background signals were marked as 0.1, and the minimum quantifiable optical signal for the QHY553M CMOS camera is 1.0. As shown in Figures 2B & D, the TOI system demonstrated a large dynamic range for IgG binding assays of the three spiked antibodies for both RBD (0.6-600 ng/mL) and S-ECD trimers (0.1-3000 ng/mL). This observation is in accordance with the fact that the homotrimers should have improved structural stability, more binding sites, and thus higher binding affinity compared with the monomers. The calibration curves constructed for the linear ranges of the IgG binding tests (Figs. 2C & E) suggested that the TOI system was able to quantify IgGs within 3-4.5 orders of magnitude (in terms of IgG concentration). To note, the calibration curves generated with S-ECD trimers exhibit a much wider dynamic range than those generated with RBDs (Figure S5). Even with the 3 orders of magnitude dynamic range for RBDs, the TOI system still outperforms traditional plate-based ELISA in quantification capability, as ELISA typically offers only 1.5 orders of magnitude in its dynamic range.

**Figure. 2.**
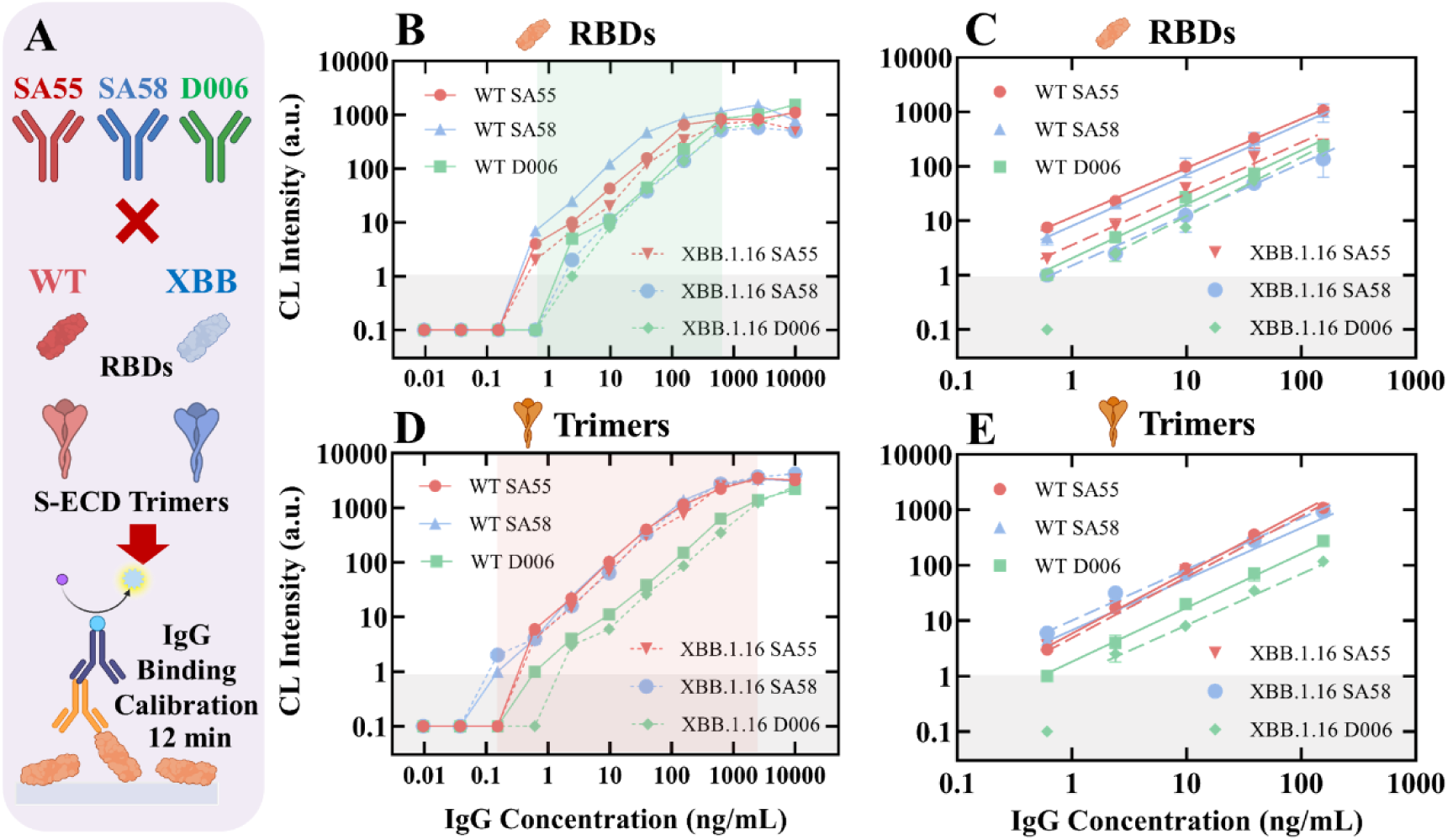
SARS-CoV-2 monoclonal antibody calibrations using TOI system. (A) Concept illustration for the IgG binding assays. Three monoclonal antibodies (SA55, SA58 and D006) were spiked for evaluation. (B&D) Large dynamic range calibration curves established with the three monoclonal antibodies that bind to SARS-CoV-2 RBD and S-ECD trimers. The linear dynamic ranges against RBDs and S-ECD trimers are 0.6-600 ng/mL and 0.1-3000 ng/mL, respectively. (C&E) Duplicated calibration curves obtained through linear regressions within the linear dynamic ranges. The error bars generated with RBDs and S-ECD trimers are ∼20% and 5-10%, respectively.

This broad linear dynamic range allows the assessment of antibodies in human blood samples without series dilution. With the convenient manual operation, the CV of this 12-minute IgG binding test ranges between 5-20%, which satisfies the requirements for the application. A significant downshift of the XBB RBD binding curves was observed compared with those of the WT strain, but such a phenomenon was not found for the S-ECD trimers. This might be attributed to the increased binding affinity of the trimeric proteins, which compensated for the decrease of antigen-antibody interaction between XBB RBD and spiked IgGs.

### Evaluation of the sample types and optimization of the IgG binding assays

Based on the established IgG binding assays with spiked antibodies, we further moved on to examine the performance difference of TOI using four common types of blood samples: fingertip blood, fingertip serum, venous blood, and venous serum (Figs. 3A & B). With the complicated matrix of blood samples from our 6 volunteers (Z1-Z6), the purposes of these experiments were to verify the robustness of the TOI system across all sample types, as well as determine the optimal dilution factor that provides the best signal-to-noise ratio.

**Figure. 3.**
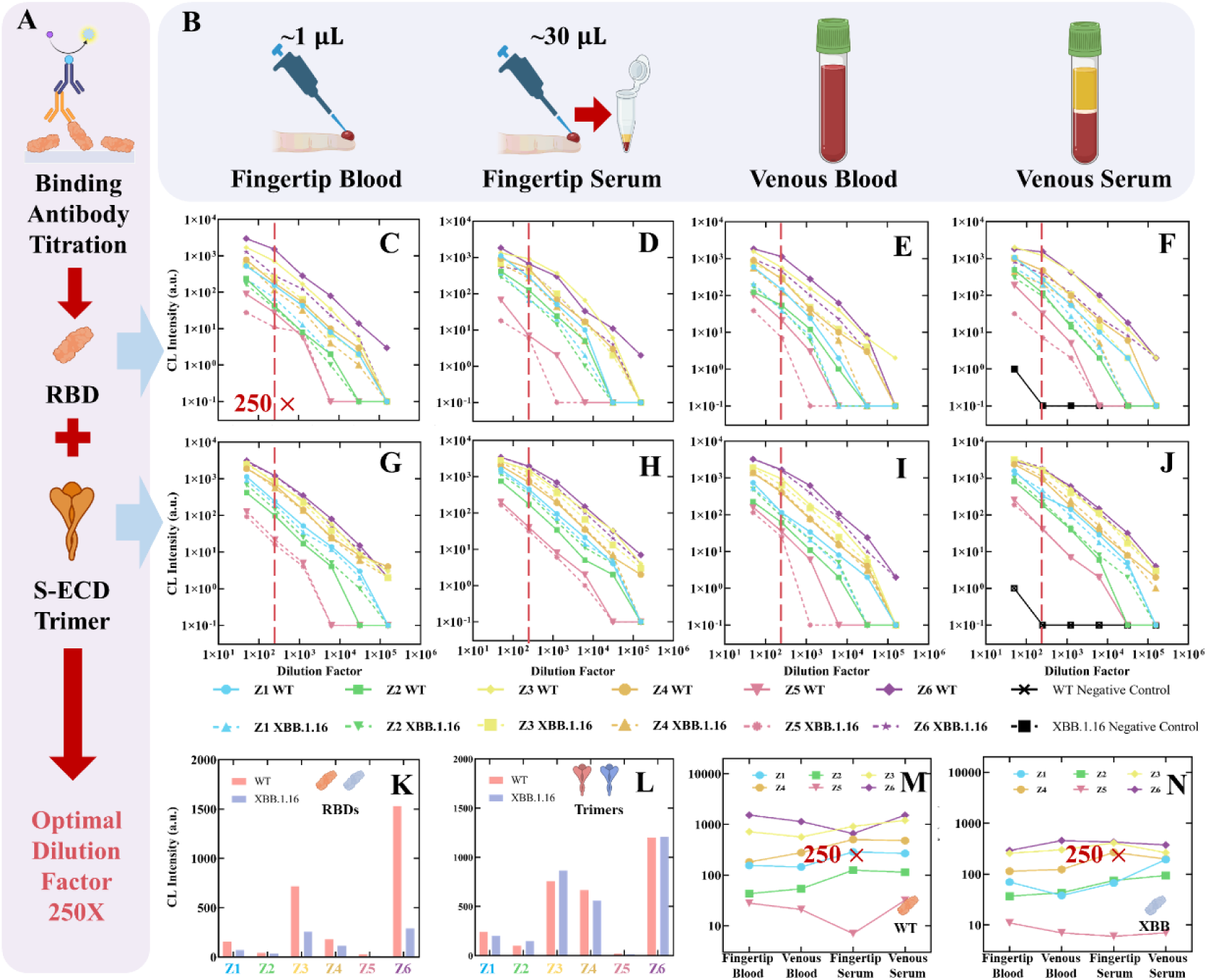
Results of the SARS-CoV-2 IgG binding titration assay, as well as the optimization of parameters. (A) Concept illustration of the IgG binding titration assay, RBDs, and S-ECD trimers from WT and XBB.1.16 strains. (B) Cross-validation of four different sample types: fingertip blood, fingertip serum, venous blood, and venous serum. (C-J) Titration curves were generated with duplicated measurements of 6 participants, Z1-Z6. For all selected antigens, 50, 250, 1250, 6250, 31260, and 156250 fold dilutions of the four sample types were verified. (5 × serial dilution) (K-L) Chemiluminescent intensities generated through the 250-time diluted fingertip blood samples against RBDs and S-ECD Trimers. (M-N) Similar chemiluminescent intensities generated with all four types of 250-time diluted samples against the RBDs.

As expected, the signals of all samples decreased as the dilution factor increased (Fig 3C-J), with the titration curves of the XBB antigens generally lying below their WT counterparts. Similar signal readings were found between the four sample types of the same individual, implying that fingertip blood is likely to be a valid source for antibody studies. The robustness of the TOI system was well illustrated, as the IgG titer rankings of the participants remained the same for all antigens. These measurements also identified a large diversity of the IgG levels across the population, as the CL signals differed over 10 times for Z5 and Z6 at all given dilutions. At the dilution factor of 250, all samples fall into the linear dynamic ranges of TOI (including the strongest and the weakest samples). This argument is valid for results generated with both RBDs and the S-ECD trimers. As a summary of all previous observations, a single dilution at 250 × should be enough for quantitative evaluation of SARS-CoV-2 binding IgG level. In other words, the traditional “titration curve” assays are no longer necessary for TOI-based immuno-evaluation. To better quantify the immuno-protection efficacy at the population level while maintaining the distinguishable signals of the background, 250 × dilution of fingertip blood will be used. Based on the triplicated measurement results presented in Fig. S6, the reproducibility of fingertip blood binding antibody assays at 250 × dilutions was remarkably good, indicating that a single-point measurement should be capable of evaluating the IgG binding quantitatively.

We further made a detailed comparison between the signal intensities generated through the 250 × diluted fingertip blood samples against RBDs and S-ECD trimers (Figs. 3K-L), as well as the CL intensities generated with all four types of diluted samples against the RBDs (Figs. 3M & N). At this dilution level, a strong preference towards WT RBD could be observed, which is generally in accordance with the vaccination and infection history of the participants. Thus, compared to the outcomes of trimers, results generated with RBD monomers will lead to a better knowledge of antibody binding specificity. Taken together, these experiments suggest that 250 × dilutions of fingertip blood could achieve a satisfactory resolution for the immuno-protection evaluation with the TOI system while being convenient and cost-efficient.

## Discussion and Conclusion

In this study, we developed a tip optofluidic immunoassay (TOI) system that can rapidly quantify one’s binding IgG level against multiple SARS-CoV-2 strains, using 1 μL of fingertip blood. Benefitted from the high performance of the microfluidic immuno-reactors and the chemiluminescent imaging station, TOI was able to achieve a lower limit of detection (LLOD) at 0.1 ng/mL (for IgG binding) and a dynamic range spanning 3-4.5 orders of magnitude, with an exceptionally high signal-to-noise ratio (∼10000), thus setting a new benchmark for convenient COVID-19 antibody assays.

During comparative analysis, we tested different types of blood samples with a series of dilution factors using the TOI system. Similar and comparable results were obtained from all types of samples, but in practice, fingertip serum was found to be the least favorable sample type. Due to the relatively large volume requirement (∼30 μL), we experienced difficulties in collecting fingertip serum from volunteers. Comparatively, 1 μL fingertip whole blood was much easier to collect and enough for all types of aforementioned assays with the TOI system.

The results generated with fingertip blood closely mirrored those obtained from traditional venous serum and whole blood samples, validating the reliability of fingertip blood as an equivalent source for antibody testing. Undoubtedly, fingertip blood-based assays can be more practical and less invasive, which is particularly beneficial for COVID-19 antibody testing.

During the binding specificity tests against SARS-CoV-2 variants, different spike protein constructs, receptor-binding domains (RBDs), and spike ectodomain (S-ECD) homotrimers were utilized. Our findings suggest that RBDs may offer a slight advantage in determining one’s SARS-CoV-2 antibody response specificity over the S-ECD homotrimers. However, it should be noted that assays using RBDs tend to have a narrower dynamic range compared to those using S-ECD homotrimers. We also identified that a 250 × dilution is optimal for RBD-based IgG binding assays, effectively distinguishing samples with higher and lower antibody abundance within the linear dynamic range of assays. This optimization is crucial for accurate and effective assessment of antibody-based immuno-protection at individual and population levels.

Beyond the direct findings presented in this work, our research extends into a comprehensive large-scale study of IgG levels in healthy volunteers. Additionally, we have renovated our rapid in-vitro inhibition assay (RIVIA), adapting it for 10 times more sensitive and efficient evaluation of SARS-CoV-2 neutralizing antibody levels with fingertip blood samples (RIVIA 2.0). These parallel efforts not only augment our understanding of the immune landscape in response to SARS-CoV-2 but also demonstrate the versatile application of our TOI platform in future vaccine development and infectious disease control. Detailed results and analyses of these extended studies will be thoroughly presented in our forthcoming official publication.

## Data Availability

All data produced in the present study are available upon reasonable request to the authors.

## Acknowledgements

This study was supported by the Shenzhen Institute of Advanced Technology, Chinese Academy of Science, and The Key Laboratory of Biomedical Imaging Science and System, Chinese Academy of Sciences. This study was also funded by the Guangdong Province Young Innovative Talents Project (2022KQNCX067), Shenzhen Basic Research Project of Natural Science Foundation (JCYJ20230808105701004), and Shenzhen Science and Technology Innovations Committee (JSGGZD20220822095200001).

The authors thank Dr. Jie Zhang and other colleagues from Sino Biological for kind support in biological reagents. The authors thank Dr. Zhengting Zou and Dr. Hongjiu Zhang from Institute of Zoology, Chinese Academy of Science for support in assay development. The authors thank Mr. Yangang Huang from Beijing 4.0 Industrial Technology Co., Ltd. for the support in microfluidic immuno-reactor fabrication. The authors also thank Dr. Yunlong Cao from Peking University for his kind support and thoughtful discussion.

