## Supplementary Information for "A Tip Optofluidic Immunoassay System for COVID-19 Immuno-protection Evaluation Using Fingertip Blood"

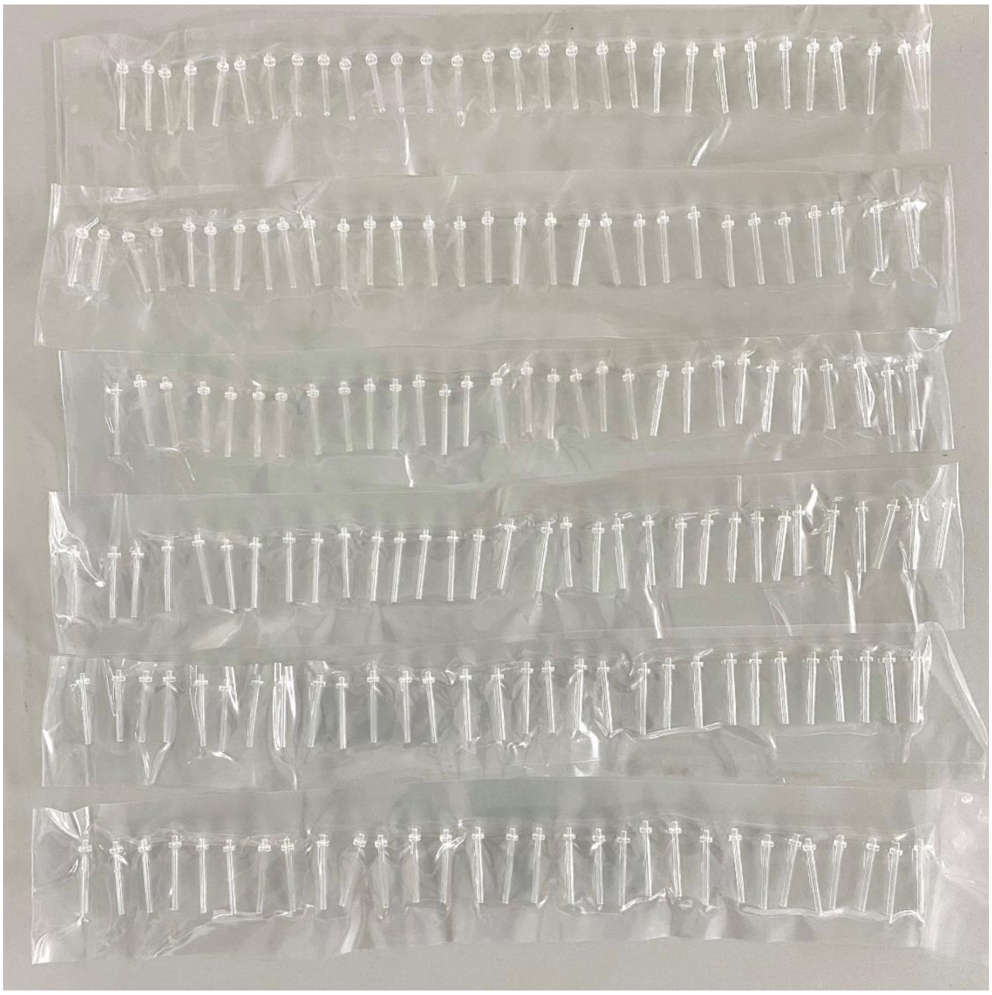

**Figure. S1.** Microfluidic immuno-reactors used in the TOI system. Fabricated through industrial-grade microscale injection molding, the batch size of the microfluidic immuno-reactor can reach 10,000 currently, with a defective rate lower than 0.1%.

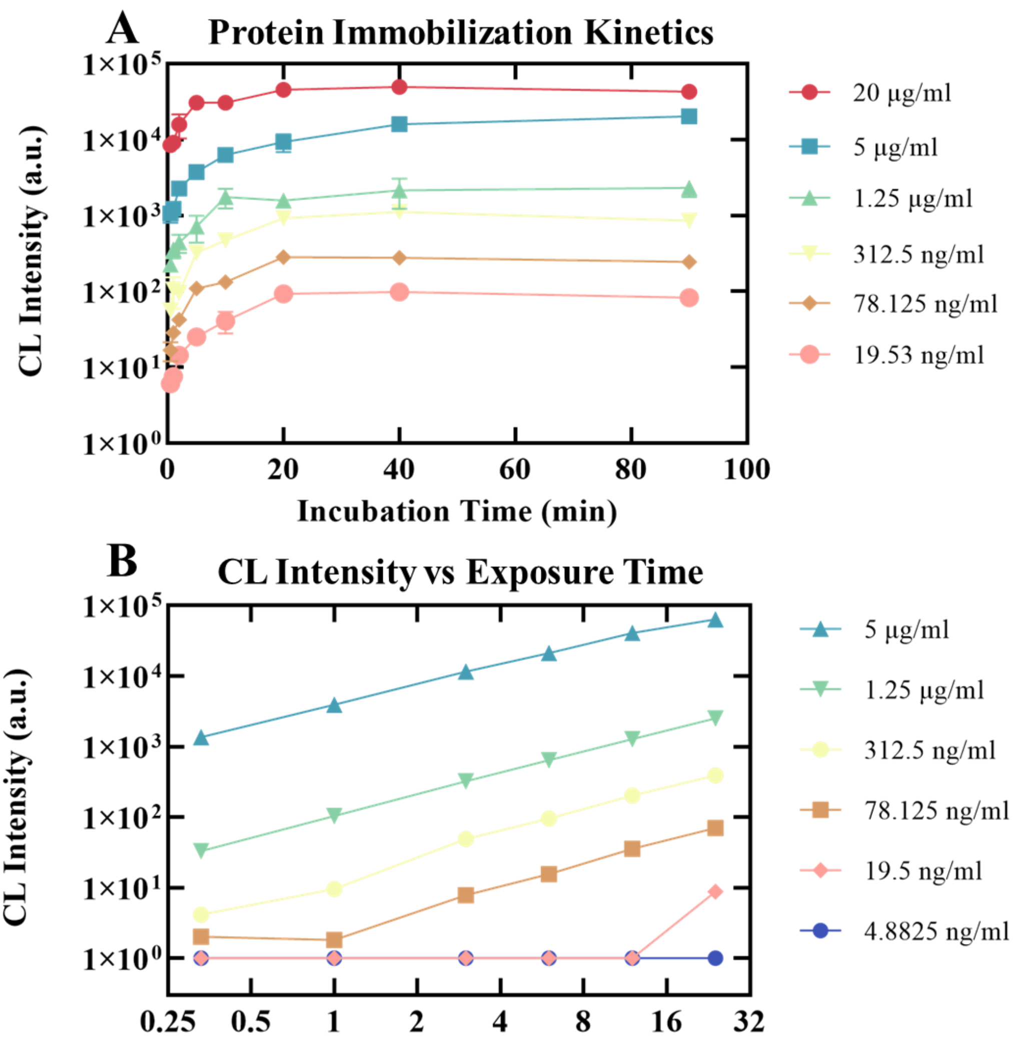


**Figure. S2.** Performance evaluation of the TOI system. (A) The protein immobilization kinetics of the microfluidic immuno-reactors. For typical proteins such as IgG, the physical adsorption generally saturates after 20-40 minutes, significantly faster than conventional immunoassay reactors like 96-well plates. (B) Sensing performance of the luminol chemiluminescent signal detected using a QHY533M CMOS camera. Nothermal noise was detected even with 12 seconds of exposure time (marked as 0.1 on the graphs), thanks to the high performance of the semiconductor cooling system in the camera. The minimum detectable optical signal intensity of the camera was 1.0, and the linear dynamic range was about 60,000 (gain = 70), covering 4.5-5 orders of magnitude in optical signal intensity.

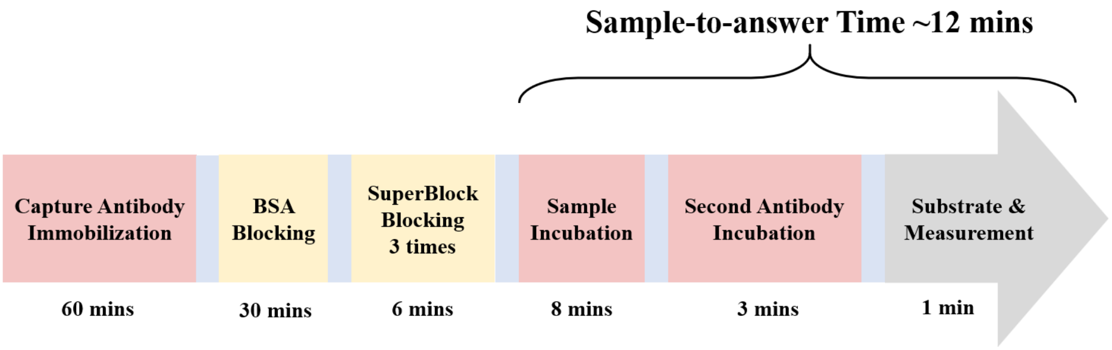


**Figure. S3.** Graphical illustration for the protocol of the IgG binding assays. The total sample-to-answer time is approximately 12 minutes. This protocol is used for both the RBD group and the S-ECD trimer group. The light blue block after each incubation step indicates a quick rinsing with PBST.

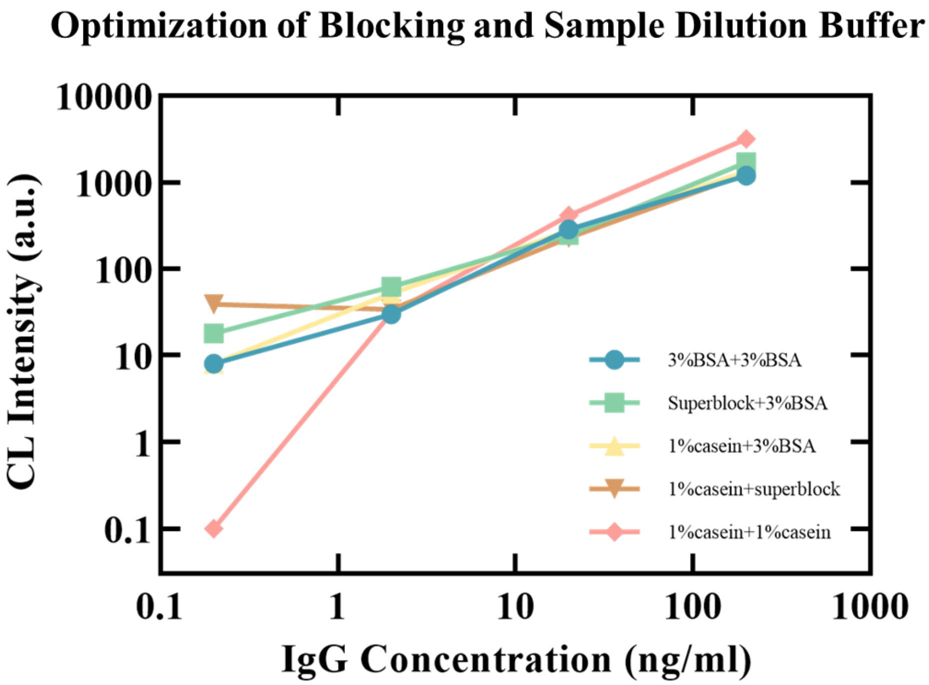


**Figure. S4.** Optimization of the blocking buffer and the sample dilution buffer. All assays were conducted according to the protocol described in Figure S3, with SARS-CoV-2 WT RBD as the target antigen and SA55 antibody as the analyte. Using 1% casein in PBS for both immuno-reactor blocking and sample/reagent dilution significantly reduced the background noise level to near zero (indicated as 0.1 on the plot).

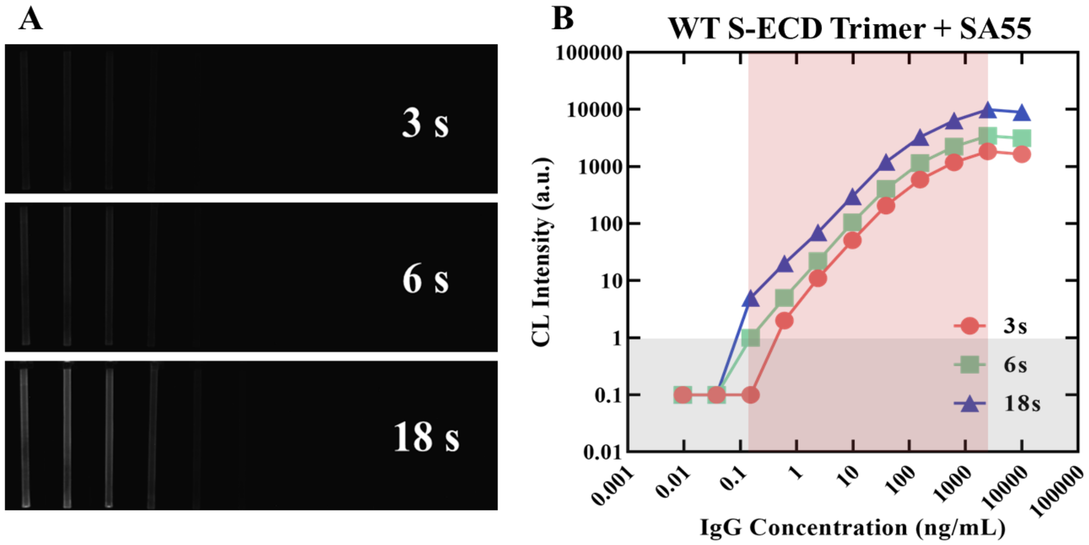


**Figure. S5.** Visual demonstration of the IgG binding assay performance. All results were generated with SARS-CoV-2 WT S-ECD trimer as the target antigen and SA55 antibody as the model analyte. (A) Chemiluminescent images collected at different exposure times (3s 6s and 18s respectively). (B) Quantification of the chemiluminescent signals from the images. An exposure time of 6s is enough for the generation of a measurable signal, even with only 0.1 ng/mL of SA55. The dynamic range of this IgG binding assay covers 0.1-3000 ng/mL (~4.5 orders of magnitude), and the weakest measurable signal intensity was 1.0 (above the grey-shaded area). Upon optimization, the signal-to-noise ratio for the IgG binding assays could reach >10000.

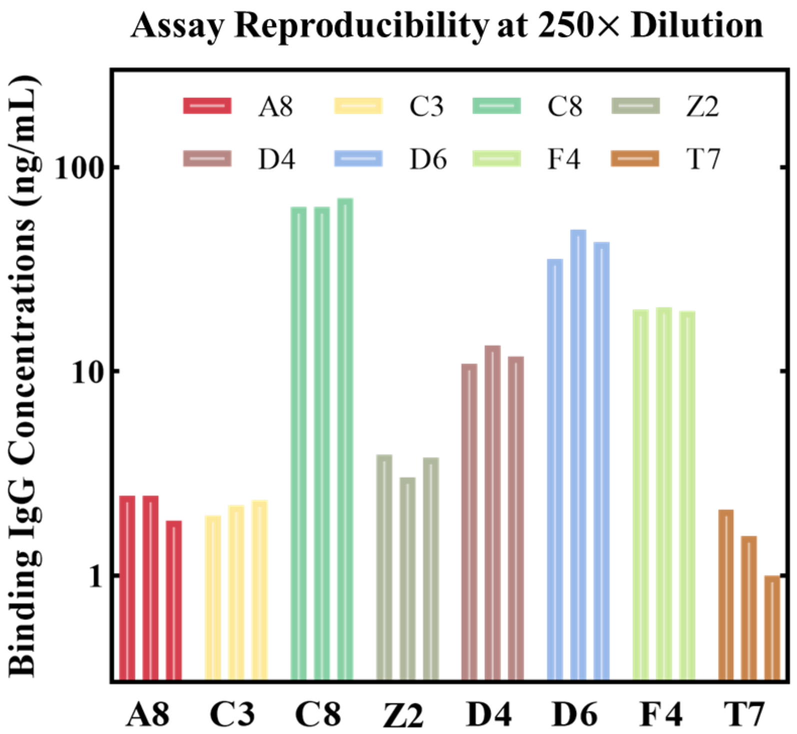


**Figure. S6.** Triplicate measurements of typical IgG binding assays using SARS-CoV-2 WT RBD as the target antigen and 250 × diluted fingertip blood as the sample. The coefficient of variation (CV) for these assays ranged from 5% to 20%, suggesting that a single-point measurement is sufficient for quantitative IgG binding evaluation
